# ASSOCIATION BETWEEN ETHNICITY AND SEVERE COVID-19 DISEASE: A SYSTEMATIC REVIEW AND META-ANALYSIS

**DOI:** 10.1101/2020.08.12.20157271

**Authors:** Antony Raharja, Alice Tamara, Li Teng Kok

**Affiliations:** Guy’s and St Thomas’ NHS Foundation Trust, London, UK; Faculty of Medicine, Universitas Indonesia, Jakarta, Indonesia

## Abstract

**Background:** Multiple reports suggest a disproportionate impact of Covid-19 on ethnic minorities. Whether ethnicity is an independent risk factor for severe Covid-19 disease is unclear.

**Purpose:** Review the association between ethnicity and poor outcomes including all-cause mortality, hospitalisation, critical care admission, respiratory and kidney failure.

**Data Sources:** MEDLINE, EMBASE, Cochrane COVID-19 Study Register, WHO COVID-19 Global Research Database up to 15/06/2020, and preprint servers. No language restriction.

**Study Selection:** All studies providing ethnicity-aggregated data on the pre-specified outcomes, except case reports or interventional trials.

**Data Extraction:** Pairs of investigators independently extracted data, assessed risk of bias using Newcastle-Ottawa scale (NOS), and rated certainty of evidence following GRADE framework.

**Data Synthesis:** Seventy-two articles (59 cohort studies with 17,950,989 participants; 13 ecological studies; 54 US-based and 15 UK-based; 41 peer-reviewed) were included for systematic review and 45 for meta-analyses. Risk of bias was low, with median NOS 7 of 9 (interquartile range 6-8). In the unadjusted analyses, compared to white ethnicity, all-cause mortality risk was similar in Black (RR:0.96 [95%CI: 0.83-1.08]), Asian (RR:0.99 [0.85-1.16]) but reduced in Hispanic ethnicity (RR:0.69 [0.57-0.84]). Age and sex-adjusted-risks were significantly elevated for Black (HR:1.38 [1.09-1.75]) and Asian (HR:1.42 [1.15-1.75]), but not for Hispanic (RR:1.14 [0.93-1.40]). Further adjusting for comorbidities attenuated these association to non-significance; Black (HR:0.95 [0.72-1.25]); Asian (HR:1.17 [0.84-1.63]); Hispanic (HR:0.94 [0.63-1.44]). Similar results were observed for other outcomes. In subgroup analysis, there was a trend towards greater disparity in outcomes for UK ethnic minorities, especially hospitalisation risk.

**Limitations:** Paucity of evidence on native ethnic groups, and studies outside the US and UK.

**Conclusions:** Currently available evidence cannot confirm ethnicity as an independent risk factor for severe Covid-19 illness, but indicates that disparity may be partially attributed to greater burden of comorbidities.

**Registration:** PROSPERO, CRD42020188421

**Funding source:** none

## INTRODUCTION

Up to 85% individuals affected by SARS-CoV-2 are asymptomatic or have mild illness, 15% require oxygen and 5% critically ill requiring intensive care unit (ICU) admission (1). Several risk factors such as age, male sex, and comorbidities have been shown to correlate with more severe disease (2). Understanding the demographic risk factors for severe disease is of paramount importance not only to inform clinical practice, but also to inform risk stratification at workplace and focus public health efforts at protecting vulnerable groups and those most affected by Covid-19 (3).

Initial reports hinted at the overrepresentation of ethnic minority groups in Covid-19 deaths and ICU admissions (4,5). Previous systematic review by Pan *et al*. highlighted the lack of ethnicity reporting in the literature; Nevertheless, it found consistent evidence of greater infection rates in ethnic minorities, but noted conflicting evidence on mortality (6). The association between ethnicity and Covid-19 outcomes may be riddled with confounding factors such as age, sex, comorbidities and socioeconomic factors (7). As such, considerations of covariates are necessary to determine if ethnicity is truly an independent risk factor for poor outcomes in Covid-19.

This systematic review and meta-analysis evaluates the association between ethnicity and poor outcomes (mortality, hospitalisation, ICU admission, advanced respiratory support, and kidney failure) in patients with laboratory-confirmed SARS-CoV-2.

## METHOD

This study was conducted according to the Preferred Reporting Items for Systematic Review and Meta-Analysis (PRISMA) Statement (8). The protocol of this review was registered in PROSPERO database, CRD42020188421.

### Data Sources and Searches

A literature search of databases was conducted on 31^st^ May 2020, and later updated on 15^th^ June 2020. Databases used were Ovid MEDLINE, Ovid EMBASE, Cochrane COVID-19 Study Register, and World Health Organization (WHO) COVID-19 Global Research Database. Search terms were “SARS-CoV-2 OR Covid-19 OR novel coronavirus” and “ethnic OR race OR minority group OR demography”. The *Lancet, BMJ*, and *JAMA* were reviewed for any relevant articles. Preprint servers (MedRxiv and BioRxiv) were searched for non-peer-reviewed preprint articles. References of included studies and a related systematic review were screened for relevant articles (6). Citation tracking was carried out using Google Scholar on 24 June 2020 to identify relevant articles which cited any of the included studies. No language restrictions were applied. All steps were carried out by pairs of independent reviewers (AT, LK, AR), and a third reviewer arbitrated for cases without consensus. Full details of search strategies are described in **Online Supplementary Appendix S2**.

### Study Selection

We included studies that reported associations between ethnicity and any of the prespecified outcomes indicative of severe Covid-19 disease in laboratory-confirmed SARS-CoV-2 patients. Primary outcome is all-cause mortality. Secondary outcomes are hospitalisation, critical care admission, advanced respiratory support requirement (such as invasive mechanical ventilation (IMV), extracorporeal membrane oxygenation (ECMO), and acute kidney injury (any severity or the need for acute renal replacement therapy). In particular, we excluded studies that reported on infection rates alone. Interventional trials, case reports, commentaries and articles from news media were excluded.

### Data Extraction and Quality Assessment

A custom spreadsheet was developed and piloted by AR and AT prior to use for data extraction; this recorded study characteristics (name, date of publication status), aim, location, setting, participant characteristics including age, sex, ethnicity (White, black, Asian, Hispanic, mixed, or missing data), body mass index, smoking status, comorbidities, and whether the study collected any data on socioeconomic factors. Raw outcome data were extracted, along with any adjusted and unadjusted risk estimates (hazard ratios (HR), relative risks (RR) and odds ratios (OR)).

The Newcastle-Ottawa Scale (NOS) and the modified NOS by Modesti were used to assess risk of bias for longitudinal cohort studies and ecological studies respectively (9,10). A maximum score of 9 is possible, with 7 and above being regarded as low risk of bias. Pairs of investigators (AR, AT and LK) independently carried out data extraction and assessed risk of bias. A third investigator resolved any disagreement through discussion. Details of the data extraction form and NOS can be found in the **Online Supplementary Appendix S3 and S4**.

GRADE framework was used to assess the quality of evidence, and certainty in the adjusted risk estimates for each ethnicity-outcome association (11). Observational studies provided high quality evidence for prognostic factors, and are down-rated for risk of bias, inconsistency, imprecision, indirectness, publication bias, or up-rated for strong evidence of association (RR>2 or <0.5) (12).

### Data Synthesis and Analysis

Meta-analysis was carried out if two or more longitudinal cohort studies compared risk of outcomes in Black, Asian or Hispanic ethnic group with white participants (reference group) for each outcome. When multiple articles studied the same patient cohort, we used the articles reporting largest number of events. Ethnicity-outcome associations in COVID-19 patients were assessed using DerSimonian-Laird random-effects meta-analyses in the R package *meta*, as we expect heterogeneity across prognostic studies. If the risk estimates were not reported but raw data available, we calculated RRs and 95% confidence intervals for cohort and single arms of case-controlled studies. RRs were used as they do not overestimate risks, and outcomes were not rare in most studies (13). ORs were converted to RRs using Zhang and Yu formula, as developed in R *(orsk)* for the purpose of meta-analysis if outcome was >10% (14). OR was assumed to approximate RR if the outcome in the study was rare.

Meta-analyses of unadjusted and adjusted risk estimates were carried out separately for each ethnicity-outcome association, with pooled log(HR) and log(RR) calculated separately (15,16). We present two sets of adjusted estimates 1) adjusted for age and sex, 2) age, sex and at minimum one comorbidity. For ease of readership and interpretation, log(HR) and log(RR) were converted to HR and RR in the texts. Study heterogeneity was evaluated by *I^2^* statistic and visual inspection of the forest plots.

Reasons for heterogeneity were explored; Subgroups analyses stratified by locations (UK or US) or risks of bias (NOS<7 or ≥7) were conducted, including assessments of interaction. As a posthoc analysis, we calculated pooled risk estimates after excluding small studies (n<100). Funnel plots were examined for publication bias and evaluated for asymmetry using Begg’s rank correlation test as data from observational studies were likely to have asymmetric distribution (17). All statistical analyses were performed in RStudio (version 1.3.959).

### Role of the funding source

There was no funding source for this study. All authors had full access to all the data in the study and had final responsibility for the decision to submit for publication.

## RESULTS

The literature search retrieved 5,706 articles on the databases, including 1,043 duplicates. Additional 296 preprint articles were screened. Titles and abstract screening excluded 4,883 articles, and full-text examination excluded 45 articles. Manual searching, reference tracking and citation tracking yielded a further 27 articles. Study selection process is illustrated according to PRISMA flowchart (**Figure 1**).

**Figure 1:**
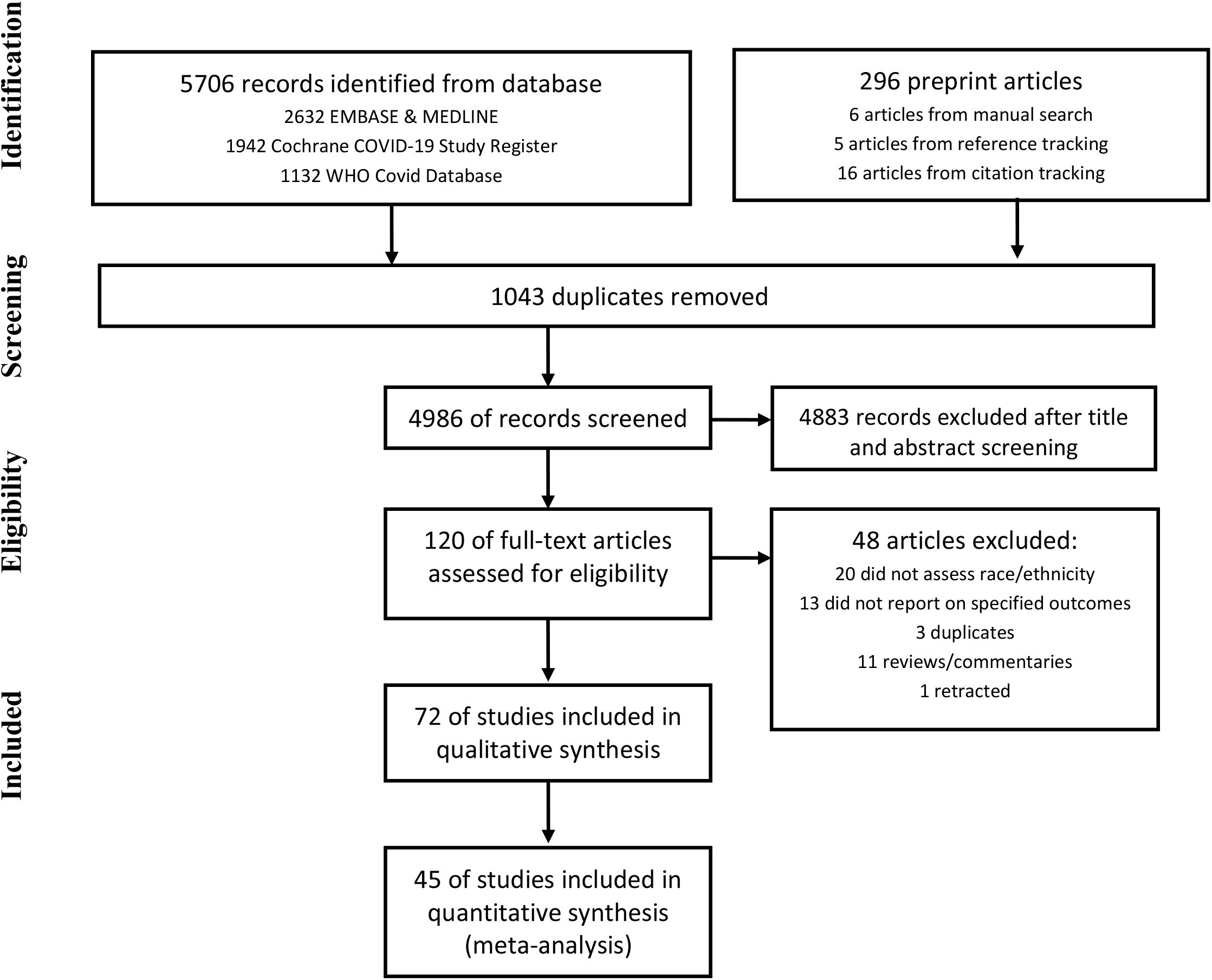
Study selection process

There were 72 articles (59 cohort and 13 ecological studies) included for qualitative synthesis; 41 (57%) studies were peer-reviewed publications. Studies were conducted in the US (54 studies), the UK (15), Brazil (one) and Israel (one). There was one multinational study based across the USA, Canada and Spain. Fifty-one studies assessed ethnic disparity in risks of mortality, 21 on hospitalisation, 18 on ICU admission, 18 on IMV, 8 on kidney failure. Adjusted analyses were carried out by 40 of 59 cohort studies and 5 of 13 ecological studies. The median NOS score was 7 (interquartile range 6-8, range 4-9). Twenty-four (33%) studies had a NOS score of less than 7, mainly due to failure to control for age or comorbidities or non-representative study sample such as pregnant women, paediatric, cancer or transplant patients (**Supplementary results S1.1-2**).

Of the 59 cohort studies, including one case-controlled study in which suitable data was extracted from one of its arms, there were 17,950,989 participants; 11,502,289 (64%) were White; 383,303 (2.1%) were Black; 1,055,396 (5.9%) were Asian; 15,439 (0.086%) were Hispanic; 4,596,081 (26%) had missing ethnicity data. Study characteristics are summarised in **Table 1**. Forty-five studies were included for meta-analysis. Summary of pooled risk estimates could be found in Table 2. A high level of heterogeneity was observed between studies. Publication bias was not detected (**Supplementary Results S4**).

**Table 1:** Summary of study characteristics

|  | Overall | Mortality | Hospitalisation | ICU admission | Kidney failure | IMV |
| --- | --- | --- | --- | --- | --- | --- |
| No of studies | 72 | 51 | 21 | 18 | 8 | 18 |
| *Peer-reviewed* | 41 | 27 | 13 | 11 | 4 | 14 |
| Study location |  |  |  |  |  |  |
| *UK* | 15 | 11 | 5 | 5 | 1 | 1 |
| *USA* | 55 | 39 | 16 | 13 | 7 | 17 |
| *Others* | 4 | 3 | 0 | 0 | 0 | 0 |
| Ecological studies | 13 | 13 | 1 | 0 | 0 | 0 |
| Cohort studies | 59 | 38 | 20 | 18 | 8 | 18 |
| No of participants in cohort studies (N) | 17,950,989* | 17,501,820 | 428,000 | 30301 | 21,999 | 16862 |
| *White, n (%)* | 11,502,289 (64) | 11,093,885 (63) | 385,113 (90) | 13611 (45) | 8,334 (38) | 6958 (41) |
| *Black* | 383,303 (2.1) | 372,020 (2.1) | 19,395 (4.5) | 9830 (32) | 8,347 (38) | 6914 (41) |
| *Asian* | 1,055,396 (5.9) | 1,045,189 (6.0) | 14,726 (3.5) | 1289 (4.3) | 1,137 (5.2) | 865 (5.1) |
| *Hispanic* | 15,439 (0.086) | 12,148 (0.069) | 6,888 (1.6%) | 2405 (7.9) | 2,007 (9.1) | 657 (3.9) |
| *Mixed or others* | 529,953 (2.9) | 514,340 (2.9) | 11,520 (3.0) | 1873 (6.2) | 2854 (13) | 941 (5.6) |
| *Missing data* | 4,596,081 (26) | 4,594,472 (26) | 834 (0.19) | 1438 (4.7) | 610 (2.8) | 718 (4.3) |
| **Newcastle-Ottawa scale points, median (IQR), range** |  |  |  |  |  |  |
| Number of studies with NOS≥ 7 points, k (%) | 48/72 (67%) | 36/51 (71%) | 13/21 (62%) | 14/18 (78%) | 8/8 (100%) | 14/18 (78%) |
| Overall | 7 (6-8), 4-9 | 7 (6-8), 4-9 | 8 (6-8), 4-9 | 8 (7.25-9), 6-9 | 8.5 (7.75-9), 7-9 | 8 (7.25-9), 6-9 |
| Selection of study groups (max 4) | 4 (3-4), 2-4 | 4 (3-4), 2-4 | 4 (3-4), 2-4 | 4 (3-4), 3-4 | 4 (3.75-4), 2-4 | 4 (3-4), 3-4 |
| Comparability of groups (max 2) | 1.5 (0-2), 0-2 | 1 (0-2), 0-2 | 2 (0.25-2), 0-2 | 2 (1.25-2), 0-2 | 2 (2-2), 0-2 | 2 (1.25-2), 0-2 |
| Outcome (max 3) | 3 (2-3), 1-3 | 3 (2-3), 1-3 | 2 (2-3), 1-3 | 3 (2-3), 2-3 | 3 (3-3), 2-3 | 3 (2.25-3), 1-3 |

\*one study was excluded as data derived from same cohort (41,42). Studies may separate ethnicity from race; in such cases, Hispanic and other races are not mutually exclusive.

**Table 2:** Summary estimates for mortality with log (hazard ratio) [95% Confidence interval], I^2^, and number of studies (k), or log (relative risk) [95% Confidence interval], I^2^, and number of studies.

| Ethnicity (white as reference) | Univariate | Multivariate† | Multivariate‡ |
| --- | --- | --- | --- |
| **Mortality:** |  |  |  |
| Black v white, log (HR) | -0.17 [-0.63; 0.30], 94, k=4 | 0.32 [0.09; 0.56]*, 94, k=5 | -0.05 [-0.33; 0.22], 79, k=4 |
| Black v white, log(RR) | -0.05 [-0.18; 0.08], 88, k=25 | 0.25 [0.03; 0.46]*, 12, k=5 | 0.06 [-0.05; 0.17], 60, k=6 |
| Asian v white, log (HR) | 0.01 [-0.31; 0.33], 82, k=4 | 0.35 [0.14; 0.56]*, 87, k=3 | 0.16 [-0.18; 0.49], 73, k=3 |
| Asian v white, log(RR) | -0.01 [-0.16; 0.15], 84, k=14 | 0.42 [0.04; 0.80]*, 0, k=2 | NA |
| Hispanic v white, log (HR) | NA | NA | -0.06 [-0.47; 0.35], 89, k=2 |
| Hispanic v white, log(RR) | -0.36 [-0.56; -0.17]*, 76, k=11 | 0.13 [-0.08; 0.34], 0, k=3 | NA |
| **Hospitalisation:** |  |  |  |
| Black v white, log(RR) | 0.52 [0.25; 0.79]*, 98, k=13 | 0.80 [0.43; 1.16]*, 92, k=5 | 0.34 [-0.07; 0.75], 95, k=4 |
| Asian v white, log(RR) | 0.13 [-0.25; 0.50], 95, k=8 | 0.15 [-0.44; 0.73], 82, k=3 | 0.04 [-0.01; 0.10], 0, k=3 |
| Hispanic v white, log(RR) | 0.00 [-0.05; 0.05], 0, k=8 | 0.40 [0.25; 0.56]*, 0, k=2 | 0.22 [0.02; 0.42]*, 75, k=3 |
| **Intensive care unit admission** |  |  |  |
| Black v white, log(RR) | 0.41 [0.11; 0.71]*, 94, k=10 | 0.33 [-0.16; 0.82], 69, k=3 | 0.27 [-0.18; 0.71], 91, k=4 |
| Asian v white, log(RR) | 0.02 [-0.67; 0.70], 71, k=4 | NA | NA |
| Hispanic v white, log(RR) | -0.12 [-0.28; 0.05], 0, k=6 | NA | -0.07 [-0.29; 0.15], 0, k=2 |
| **Invasive mechanical ventilation** |  |  |  |
| Black v white, log(RR) | 0.25 [-0.11; 0.60], 85, k=10 | 0.34 [0.12; 0.56]*, 0, k=3 | 0.21 [-0.51; 0.92], 91, k=3 |
| Asian v white, log(RR) | 0.33 [0.07; 0.59]*, 14, k=4 | 0.43 [0.16; 0.70]*, 0, k=2 | NA |
| Hispanic v white, log(RR) | -0.12 [-0.37; 0.14], 0, k=6 | NA | 0.01 [-0.17; 0.19], 0, k=2 |
| **Acute kidney injury** |  |  |  |
| Black v white, log(RR) | 0.30 [0.04; 0.56]*, 92, k=5 | NA | -0.47 [-0.12; 1.07], 95, k=2 |
| Asian v white, log(RR) | -0.17 [-0.35; 0.01], 0, k=2 | NA | NA |
| Hispanic v white, log(RR) | NA | NA | NA |

\*statistically significant; †Adjusted for age and sex; ‡Adjusted for age, sex and comorbidities; NA: data not available

**Table 3:** Quality of evidence assessment. Age, sex and comorbidities adjusted-risk estimates for each ethnicity-outcome association, and application of GRADE principles towards rating confidence in risk estimates

|  | **All-cause Mortality** | **Hospital isation** | **ICU** | **Ventilation** | **AKI or acute RRT** | Risk of bias | Inconsistency | Imprecision | Indirect ness | Publication bias | Strong association | GRADE |
| --- | --- | --- | --- | --- | --- | --- | --- | --- | --- | --- | --- | --- |
| Black | Ntrl | Ntrl | Ntrl | Ntrl | ↑ | - | - | - | - | - | - | HIGH |
| Asian | Ntrl | Ntrl | NA | NA | NA | DOWN | - | - | DOWN | - | - | LOW |
| Hispanic | Ntrl | ↑* | Ntrl | Ntrl | NA | DOWN | - | - | DOWN | - | - | LOW |

GRADE= Grading of Recommendations Assessment, Development and Evaluation; ↑\* = significant risk factor; ↑ = potential risk factor; non-significant risk ratio >1.5; Ntrl = Neutral association; risk ratio between 0.67 and 1.5; ↓ = potentially protective; non-significant risk ratio <0.67; ↓\* = significant protective; NA = not applicable

### Mortality

Fifty-one studies reported ethnicity-aggregated mortality data, including 38 cohort studies comprising 17,501,820 participants (63% White, 2.1% Black, 6.0% Asian, 0.069% Hispanic, 2.9% others, and 26% missing ethnicity data). Total sample sizes were more than 100 participants (n>100) in 26 of 28 (93%) cohort studies included in the meta-analysis.

Pooled estimates from hazard ratios and relative risks generally showed similar magnitude, direction of effect and statistical significance. In the unadjusted analyses, compared to white ethnicity, all-cause mortality risk was similar in Black (RR:0.96 [95%CI:0.83-1.08, I^2^=88, k=25), Asian (RR:0.99 [95%CI: 0.85-1.16], I^2^=84, k=14) but significantly reduced in Hispanic ethnicity (RR: 0.69 [95%CI: 0.57-0.84], I^2^=76, k=11). Age and sex-adjusted mortality risks were significantly elevated for Black (HR: 1.38 [95%CI: 1.09-1.75], *I*^2^=94, k=5) and Asian (HR: 1.42 [95%CI: 1.15-1.75], *I*^2^=87, k=3), but not for Hispanic (RR: 1.14 [95%CI: 0.93-1.40, I^2^=0, k=3). Further adjustment for comorbidities attenuated these associations, rendering associations non-significant; HR (Black): 0.95 [95%CI: 0.72-1.25], I^2^=79, k=4; HR (Asian): 1.17 [95%CI: 0.84-1.63], I^2^=73, k=3; HR (Hispanic): 0.94 [95%CI: 0.63-1.44], I^2^=89, k=2.

Subgroup analysis by location showed a consistent trend towards greater mortality risk estimates in UK ethnic minorities, but difference was not significant. Subgrouping by risk of bias did not demonstrate different effects (**Supplementary Results S2**)

Ten cohort studies, not included in meta-analysis, echoed similar findings, and did not support ethnicity as an independent risk factor for poor Covid-19 outcomes. Two studies reported non-significant difference in unadjusted mortality risk in Black (v. non-Black) (18,19). Four studies (three unadjusted, one adjusted analyses) did not find an increase in mortality risk amongst non-White ethnicity (20–23). Four studies reported lower mortality risk in Hispanic patients compared to non-Hispanic patients; two reported significant unadjusted analysis, and none reported significant age-adjusted analyses (24–27).

There were 12 US-based ecological design studies, utilising publicly-available dataset to draw an indirect association between ethnicity and Covid-19 outcomes. Eight studies illustrated that counties with greater proportion of African-Americans have higher rates of Covid-19 hospitalisation and death (28–32), even after adjusting for a combination of county-level characteristics such as age, poverty, comorbidities, healthcare access, geography (33–36). One study did not find an increased mortality risk in Asian ethnicity after adjustment for age and geography (35). Instead, a higher proportion of Asian population was a protective factor to counties (32). Three studies reported increased age-adjusted mortality in the Hispanic population (32,35–37). One study noted that ethnic segregation correlated with risks of Covid-19 death (38). One UK study showed that Black African, Black Caribbean, Pakistani, Bangladeshi and Indian ethnic groups had higher age-standardised and geography-adjusted mortality ratio (39).

### Hospitalisation

Twenty-one studies assessed hospitalisation risk in different ethnic groups. There were 20 cohort studies comprising 428,000 patients (90% White, 4.5% Black, 3.4% Asian, 1.6% Hispanic, 3.0% others, and 0.19% missing ethnicity data); 14 articles were suitable for meta-analysis. Only one had a small sample size n<100.

Compared to White ethnicity, crude unadjusted risk of hospitalisation was significantly higher in Black (RR: 1.68 [95%CI: 1.28-2.20], I^2^=98, k=13), but similar for Asian (RR: 1.13 [95%CI: 0.78-1.66], I^2^=95, k=8) and Hispanic (RR: 1.00 [95%CI: 0.95-1.06], I^2^=0, k=8) ethnicity. Age and sex-adjusted risk was significantly raised in Black (RR: 2.23 [95%CI: 1.54-3.19], I^2^=92, k=5) and Hispanic (RR: 1.49 [95%CI: 1.28-1.75], I^2^=0, k=2), but not Asian (RR: 1.16 [95%CI: 0.64-2.08], I^2^=82, k=3) ethnicity. Weakening of association was noted after further adjustment for comorbidities: Black (RR: 1.40 [95%CI: 0.93-2.12], I^2^=95, k=4), Asian (RR: 1.04 [95%CI: 0.99-1.11], I^2^=0, k=3), and Hispanic (RR: 1.24 [95%CI: 1.02-1.52], I^2^=75, k=3). Five studies considered further socioeconomic factors in their analysis and showed that adjusting for socioeconomic factors could reduce the disparity in hospitalisation risk (40-44).

Subgroup analysis showed strongly significant interaction p value between UK and US subgroups. The hospitalisation risk of Black and Asian were markedly higher in UK. For Black ethnicity, RR 5.47 [95%CI: 2.51; 12.06] in UK studies v. RR 1.36 [95%CI: 1.08; 1.72] in US studies with p value of 0.0008. For Asian ethnicity, RR: 2.95 [95%CI: 1.55-5.53] in UK studies *v*. RR: 0.90 [95%CI: 0.82-1.66] in US studies with p value of 0.0003. Subgrouping by location was not possible for Hispanic ethnicity. Subgrouping by NOS did not show significant interaction, although there was a trend towards greater risk in studies with lower NOS (**Supplementary Results S2**).

Seven studies were not suitable for meta-analyses. In a descriptive unadjusted analysis, three studies reported no differences between the number of Hispanic and non-Hispanic patients (25), Black and non-Black patients (18), and between white and BAME patients (20). Despite no significant differences, ethnic minorities were still disproportionately represented in Covid-19 admissions (45,46). Two other studies reported increased adjusted-risks of hospitalisation in non-white ethnic groups in both US and UK (42,47). One New York study showed that rates of Covid-19 hospitalisations were higher in areas with greater proportion of ethnic minorities (28).

### Critical care admission

Eighteen studies assessed ethnicity as a risk factor for ICU admission, comprising 30,301 participants (45% White, 32% Black, 7.9% Asian, 7.9% Hispanic, and 4.7% with missing ethnicity data). Subgroup analyses by location or NOS scores were not possible as ten of 11 studies included in meta-analyses were US-based, and all have NOS≥7.

Compared to White ethnicity, the unadjusted risks of ICU admission were significantly raised in Black (RR: 1.51 [95%CI: 1.11-2.04], I^2^=94, k=10), similar in Asian (RR: 1.02 [95%CI: 0.51-2.22], I^2^=71, k=4), and lower in Hispanic ethnicity (RR: 0.89 [95%CI: 0.75-1.05], I^2^=0, k=6). However, pooled estimates for Asian included three US-based studies and one UK study. The largest study was UK-based and reported an increased risk for Asian ethnicity (48). The other three studies were US-based studies, relatively smaller sample sizes, showing non-significant associations.

Risk of ICU admission for Black ethnicity was attenuated to non-significant level after adjusting for age and sex (RR: 1.39 [95%CI: 0.85-2.27], I^2^=69, k=3), and further in fully adjusted analysis (RR: 1.31 [95%CI: 0.84-2.03], I^2^=91, k=4). There was inadequate data for meta-analysis for Asian ethnicity; one study reported significantly increased age and sex-adjusted risk of ICU Admission for Asian ethnicity (49). There were two studies reporting fully adjusted analysis for Hispanic ethnicity, showing non-significantly lower risk of ICU admission (RR: 0.93 [95%CI: 0.74-1.16], I^2^=0, k=2).

Seven studies were not suitable for meta-analysis. Five UK-based studies reported over-representation of the BAME communities in ICU cohorts (20,21,46,50,51), with two reporting higher age adjusted-risk for BAME (20,21). On the other hand, two US-studies did not find a significant difference in risk of ICU admission between Black and non-Black study participants (18,19).

### Respiratory failure

Eighteen cohort studies comprising 16,862 participants (41% white, 41% black, 5.1% Asian, 3.9% Hispanic and 4.3% missing ethnicity data) reported ethnicity-aggregated data on the need for advanced respiratory support i.e. invasive mechanical ventilation (IMV). Thirteen studies were suitable for meta-analysis.

In unadjusted analyses, compared to White ethnicity, risk of IMV was higher in Black (RR: 1.28 [95%CI: 0.90-1.81], I^2^ = 85%, k=10) and Asian (RR: 1.39 [95%CI: 1.07-1.80], I^2^=14, k=4) but lower in Hispanic ethnicity (RR: 0.89 [95%CI: 0.69-1.15], I^2^=0, k=6). Age and sex-adjusted risks were significantly high for Black (RR: 1.40 [95%CI: 1.13-1.75], I^2^=0, k=3) and Asian ethnicity (RR: 1.54 [95%CI: 1.17-2.02], I^2^=0, k=2); There was only one study reporting non-significantly raised age-and-sex-adjusted risk of IMV in Hispanic patients (52). After full adjustment, the associations were attenuated and non-significant in Black (RR: 1.23 [95%CI: 0.61-2.51], I^2^=91, k=3) and Hispanic ethnicity (RR: 1.01 [95%CI: 0.84-1.21], I^2^=0, k=2). One study reported a non-significantly lower age, sex and comorbidity-adjusted risk of IMV in Asian (53).

Subgrouping by location was not possible as all but one study was US-based. In subgroup analysis by risk of bias, unadjusted risk of IMV for Asian and Hispanic ethnicity was reported higher by studies with low risk of bias, albeit there was no significant interaction p value between subgroups. Inclusion of studies with high risk bias did not affect the direction or the statistical significance of the overall pooled estimate, although attenuates the magnitude.

Results from studies that were not suitable for meta-analysis further reiterated that ethnicity was not associated with risk of intubation. Four studies showed that Black ethnicity was not independently associated with IMV, or a composite of IMV or death (18,19,26,54). Two studies reported similar rates of IMV in Hispanic and other ethnicities (25,26), although one study found non-significantly lower risk of IMV or death in Hispanics (54). One found non-White ethnicity to be more likely to require high-flow oxygen support or IMV, albeit this was statistically non-significant (47). We did not find any studies reporting association between ethnicity and ECMO.

### Kidney failure

Eight studies comprising 21,999 participants (38% white, 38% Black, 5.2% Asian, 9.1% Hispanic, 2.8% missing ethnicity data) investigated the association between ethnicity and acute kidney injury (AKI); all had low risk of bias and seven were US-based studies. Five studies were included in the meta-analysis.

In an unadjusted analysis, Black ethnicity was at a significantly higher risk of AKI (RR: 1.35 [95%CI: 1.04-1.76], I^2^ = 92%, k=5). Two separate studies showed that this association remained significant after adjustment of age, sex, and comorbidities, but pooled RR was non-significant (RR: 1.60 [95%CI: 0.89-2.90], *I*^2^ = 95%, k=2) most likely due to only two studies being suitable. Three studies reported higher rates of Black patients requiring acute renal replacement therapy, although none showed significant association in adjusted analysis (19,48,55).

Two studies reported lower unadjusted-risk of AKI in Asian ethnicity (RR: 0.85 [95%CI: 0.71-0.01], *I*^2^=0, k=2). One study reported non-significantly lower adjusted-risk of AKI in Asian ethnicity (56). Two studies did not find an increased unadjusted-risk of AKI in Hispanic ethnicity (25,57). There were no studies reporting adjusted risk of AKI for Hispanic ethnicities.

### Quality assessment

The level of evidence was high for Black ethnicity, but low for both Asian and Hispanic ethnicities. The certainty in the risk estimates for Asian and Hispanic was down-rated for risk of bias and indirectness due to relatively low number of studies providing age, sex and comorbidity-adjusted association, and potential differences between study participants and target population. Detailed assessment is described in the Supplementary Results S5.

## DISCUSSION

This systematic review and meta-analysis of currently available evidence did not confirm ethnicity as an independent risk factor for poor outcomes in Covid-19 patients. Analyses of step-wise adjustments for covariates underlined important factors confounding ethnicity as a risk factor.

Interpretation of ethnicity-stratified data requires considerations of traditional risk factors for covid-19. Unadjusted risk ratios could quantify crude risk disparities between different ethnicities, but may misrepresent the true association. For instance, lower unadjusted mortality risk in Hispanic masks the fatality seen in younger Hispanic group. The meta-analysis demonstrates significantly elevated age and sex adjusted-risks across several outcome measures. The attenuation of these estimates by further adjustment for comorbidities indicates that disparities could be partially attributed to a greater burden of comorbidities in ethnic minority groups. Socioeconomic factors have also been suggested to contribute to this disparity, albeit our review underlined paucity of evidence.

To our knowledge, this is the first meta-analysis quantifying the association between ethnicity and covid-19 outcomes. Our finding is in keeping with findings from pandemic influenza refuting ethnicity as a risk factor (58). Nonetheless, the evidence is consistent on the disproportionate representation of ethnic minorities in covid-19 mortality and morbidity. In fact, racial disparities during a pandemic outbreak appears to be a recurring phenomenon (59). As such, efforts to reduce disparities should be encouraged.

Substantial heterogeneity is attributed to difference in magnitude rather than the direction of effect. Methodological differences such as i.e. dissimilar combinations of comorbidities adjusted for also contributed to overall heterogeneity, but has not necessarily render our findings less useful. Clinical heterogeneity is also expected in risk estimates for Asians since Asian ethnicity is not a homogenous group, consisting of individuals from widely diverse origins such as Indian, Pakistani, Bangladeshi, Chinese and others. Subgrouping by location aims to provide context-specific and clinically useful risk estimates, whilst sacrificing precision for general applicability in public health policy decision-making. Therefore, we argue that a high level of heterogeneity has not limited the usefulness of the meta-analyses. Nevertheless, for this reason, we down-rate certainty of risk estimates for Asian and Hispanic ethnicity.

This study has clinical and public health implications. Findings from this review should inform decisions regarding risk stratification at work, shielding advice, future allocation of vaccinations. Given the low-to-high quality evidence indicating that ethnicity is not an independent risk factor, covid-19 risk assessment should only consider ethnicity in conjunction with other risk factors such as age or comorbidities. Public campaigns need to target ethnic minority groups, who appear to be overrepresented in multiple cohorts. More importantly, public health planning is required to tackle the underlying reasons for racial health disparities such as implementing measures to reduce the burden of comorbidities in ethnic minorities.

Our study has several strengths. Search strategy was comprehensive, covering large number of published peer-reviewed and preprint articles. A large number of studies was included in the meta-analysis with only several small studies sizes (n<100); Omission of these studies in post-hoc analysis did not affect the direction and statistical significance of analyses. Multiple indicators of poor outcomes i.e. mortality, hospitalisation, ICU admission, intubation, and kidney failure were considered. Taken together, the balance of evidence weighs against ethnicity as an independent risk factor for poor outcomes in patients with laboratory-confirmed SARS-CoV-2 infection. We did not investigate whether ethnicity is associated with risk of SARS-CoV-2 infection as this relates to incidence rather than severity. Besides, this been explored in previous systematic review (6). Lastly, separate pooling of unadjusted and adjusted risk-estimates improves our understanding of ethnicity-outcome association.

Our study is limited by certain gaps in currently available evidence: substantial number of ethnicity data missing in studies; limited number of articles investigated the significance of socioeconomic factors as a confounding factor to ethnicity, and area-level measurements were used rather than patient-level data; Relative paucity of evidence assessing adjusted-risks in Hispanic and Asian groups, and minimal articles studying native populations. There was a minimal involvement of paediatric patients in our meta-analysis, and so findings should not be extrapolated to paediatric patients. Given reports of association between SARS-CoV-2 and Kawasaki-like disease, this area needs to be explore further.

Despite these limitations, our rigorous study updates current evidence on the association between ethnicity and poor covid-19 outcomes, and identifies gaps in evidence that future studies can work towards.

## Data Availability

Full datasets can be obtained from the corresponding
author at

## Financial support

none.

## Disclosures

All authors have disclosed no conflict of interest.

## Reproducible Research Statement

*Study protocol:* see Methods and supplement. *Statistical code:* See Methods *Data set:* all included studies are publicly available, additional data are available upon request from corresponding author.

## Notes

### Competing Interest Statement

The authors have declared no competing interest.

### Funding Statement

No funding obtained

### Author Declarations

Ethical approval was not required for this research.

